# Low dose prime and delayed boost can improve COVID-19 vaccine efficacies by increasing B cell selection stringency in germinal centres

**DOI:** 10.1101/2021.09.08.21263248

**Authors:** Amar K. Garg, Soumya Mittal, Pranesh Padmanabhan, Rajat Desikan, Narendra M. Dixit

## Abstract

The efficacy of COVID-19 vaccines appears to depend in complex ways on the vaccine dosage and the interval between the prime and boost doses. Unexpectedly, lower dose prime and longer prime-boost intervals have yielded higher efficacies in clinical trials. To elucidate the origins of these effects, we developed a stochastic simulation model of the germinal centre (GC) reaction and predicted the antibody responses elicited by different vaccination protocols. The simulations predicted that a lower dose prime could increase the selection stringency in GCs due to reduced antigen availability, resulting in the selection of GC B cells with higher affinities for the target antigen. The boost could relax this selection stringency and allow the expansion of the higher affinity GC B cells selected, improving the overall response. With a longer dosing interval, the decay in the antigen with time following the prime could further increase the selection stringency, amplifying this effect. The effect remained in our simulations even when new GCs following the boost had to be seeded by memory B cells formed following the prime. These predictions offer a plausible explanation of the observed paradoxical effects of dosage and dosing interval on vaccine efficacy. Tuning the selection stringency in the GCs using prime-boost dosages and dosing intervals as handles may help improve vaccine efficacies.

## INTRODUCTION

The COVID-19 pandemic continues to rage and warrants intensifying the ongoing global vaccination programs (1, 2). With limited vaccine supplies, it becomes critical to identify dosing protocols that would maximize vaccine efficacy (3, 4). With the Oxford-AstraZeneca vaccine, where dosing protocols were adjusted during the trials, data has become available of the effects of different dosages used for the prime and boost doses and of different intervals separating them on vaccine efficacy (5-8). A recent study has also examined the effects of increasing the interval beyond those in the trials (9). Intriguingly, the efficacy in preventing symptomatic infection was 63.1% when a standard dose (containing 5×10^10^ virus particles) was used for both prime and boost, whereas the efficacy was substantially higher, 80.7%, when a low dose prime (containing 2.2×10^10^ virus particles) followed by the standard dose boost was administered (5). Furthermore, the efficacy increased with the interval between the prime and boost, from 55.1% at <6 weeks to 81.3% at ≥12 weeks, when standard doses were used for both (5). Inspired by these observations, studies are examining the effects of lower dosages and increased dosing intervals with other vaccines too, specifically the Pfizer-BioNTech (10-12) and Moderna (13) vaccines. An understanding of these effects would help identify optimal dosing protocols and maximize the impact of the ongoing vaccination programs. The origins of the effects remain to be elucidated.

While the role of cellular immunity is yet to be fully elucidated (14), several studies suggest that the efficacy of currently approved COVID-19 vaccines is attributable to the neutralizing antibodies they elicit (6, 11, 15-20). The higher efficacies observed above are thus argued to be due to the improved quality and quantity of the antibodies produced by the associated dosing protocols (5, 8, 9, 11, 21). For instance, higher antibody levels were observed following the boost upon increasing the dosing interval (9, 10). In some cases, antibody-dependent cellular functions too appeared to be better with the longer intervals (21). A question that arises is how the different dosing protocols elicit antibodies of different amounts and affinities for their targets.

Antibody production following vaccination (or natural infection) occurs in germinal centers (GCs) (22, 23). GCs are temporary anatomical structures assembled in lymphoid organs where B cells are locally selected based on the ability of their receptors to bind and internalize antigen presented as immune complexes on follicular dendritic cell surfaces in the GCs. (GCs can last anywhere from a few weeks to many months (23-25).) This process, termed affinity maturation, culminates, typically in weeks, in the selection of B cells with affinities that can be several orders of magnitude higher for the target antigen than those at the start of the GC reaction (26, 27). What determines the final affinities is an important question in immunology and is yet to be resolved (28-30). Several studies have identified factors that influence affinity maturation (26, 31-37). A key factor is antigen availability within GCs–related here to the vaccine dosage and antigen half-life–elucidated first by the classic experiments of Eisen and colleagues (26): B cells compete for antigen in the GCs. Their survival depends on how much antigen they acquire, as we explain below. Thus, if antigen is scarce, the selection is stringent and leads to the survival of those B cells that have high affinity for the target antigen. This phenomenon governing the GC reaction is manifested widely, including in the effects of passive immunization following HIV infection, and can be potentially exploited by tuning antigen availability (34, 35, 38, 39).

Here, we reasoned that one way in which the effects of the different vaccination protocols could arise was from the influence the protocols had on antigen availability and hence selection stringency within GCs. Specifically, low dose prime is expected to result in low antigen availability and may lead to the selection of higher affinity B cells. The standard dose boost could then enable the expansion of these higher affinity B cells. With a larger dosing interval, the decay of antigen between doses could cause an increase in selection stringency, resulting in a similar effect. To test this hypothesis, we developed a detailed stochastic simulation model of the GC reaction. Such simulation models have been shown to mimic the GC reaction faithfully and have helped resolve confounding experimental observations and predict optimal vaccination protocols (34-36, 39-44).

## RESULTS

### Stochastic simulation model of the GC reaction post COVID-19 vaccination

We present an overview of the model here (Fig. 1); details are in Methods. We considered individuals who were not previously infected and were administered COVID-19 vaccines. We focused on the GC reaction in such individuals. The simulation, building on previous protocols (35, 36, 39, 40, 42), considered and modelled events within an individual GC. The GC reaction is initiated by B cells of low affinity for a target, non-mutating antigen. The target could be a portion of or the entire spike protein of SARS-CoV-2. We simulated the ensuing affinity maturation process using a discrete generation, Wright-Fisher-type, formalism (36, 39). The GC is divided into a light zone and a dark zone (Fig. 1A). The antigen is presented in the light zone and is represented as a bit-string of *L* amino acids. Each B cell is identified by its B cell receptor (BCR), which is also represented as a bit-string of *L* amino acids. The affinity of a B cell for the antigen is determined by the extent of the match between the BCR and antigen sequences (39, 42), defined here using *ε. ε*=0 if the two sequences are completely distinct, whereas *ε*=*L* if they are identical. The higher the *ε*, the higher is the affinity. In each generation, we let each B cell have an average of η attempts to acquire antigen. η thus serves as a surrogate of antigen availability in the GC (39). The probability with which a B cell acquires antigen in each attempt is set proportional to its affinity for the antigen (39). If a B cell fails to acquire a minimum amount of antigen, it is assumed to undergo apoptosis (31), and is eliminated. The surviving B cells then compete for help from T follicular helper (T_fh_) cells. The probability that a B cell receives such help is set proportional to the amount of antigen it has acquired relative to that of the other B cells in the generation (39). B cells that do not succeed in receiving T_fh_ help are again assumed to undergo apoptosis (31). Among the surviving B cells, following previous studies (39), we let 5% exit the GC, become plasma cells, and produce antibodies; 5% exit and become memory B cells; and 90% migrate to the dark zone, where they proliferate and mutate their BCR genes and return to the light zone (39, 43). The latter B cells form the pool for the next generation of the GC reaction. The antibodies produced by plasma cells can feedback into the GC and, by displacing lower affinity antibodies in the immune complexes or by masking antigen, tend to increase the selection stringency (35, 39, 45).

**Figure 1.**
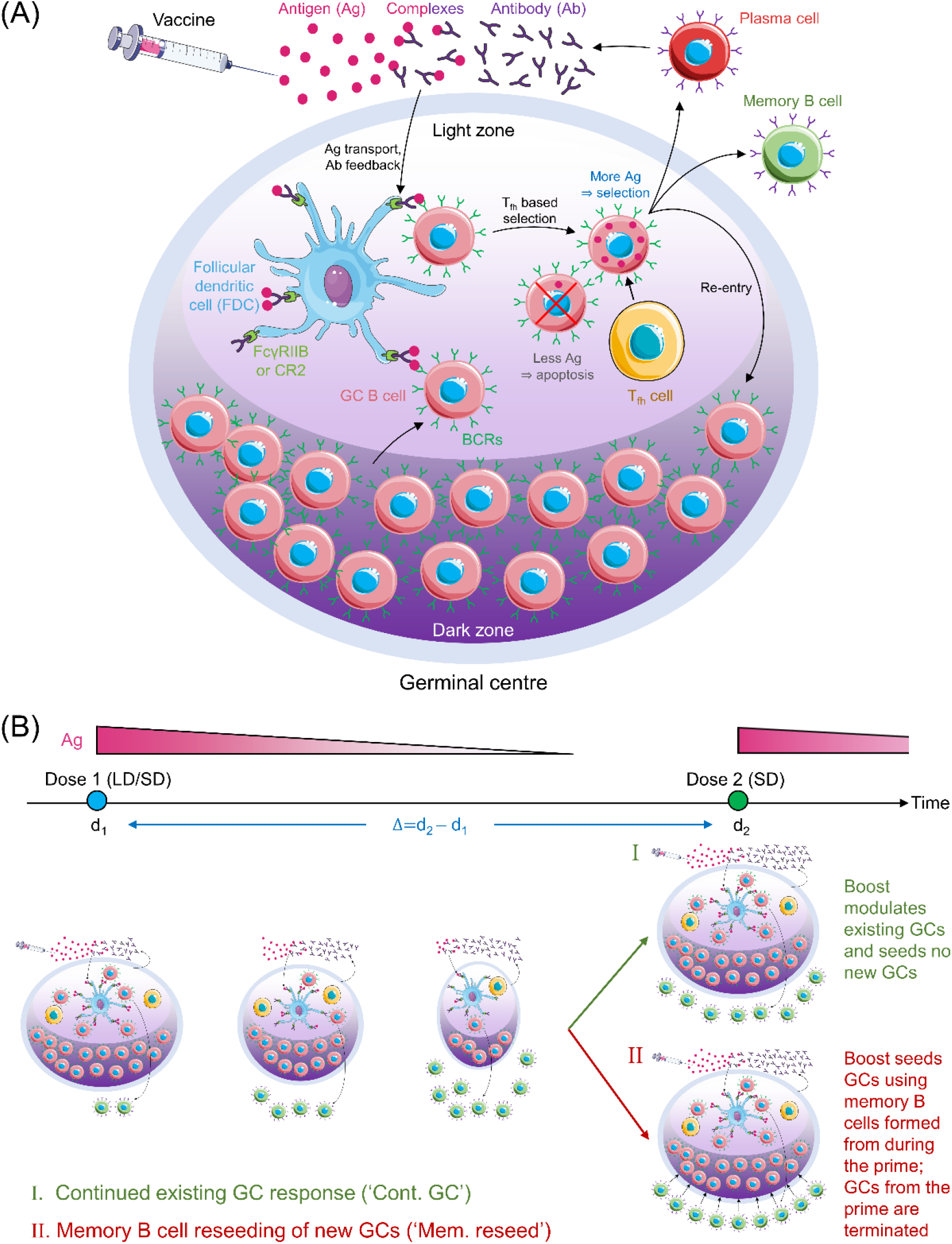
Schematic of the GC reaction model post vaccination. (A) The GC reaction. The antigen from the vaccine enters the GC complexed to antibodies and is presented in the light zone on the surfaces of follicular dendritic cells attached to FcγRIIB or CR2 receptors. GC B cells acquire antigen with a probability proportional to their affinity for the antigen. They then receive help from T follicular helper cells with a probability dependent on the relative amount of antigen they acquired. Cells that fail to acquire antigen or receive the latter help die. Cells that succeed can exit the GC to become plasma cells and secrete antibodies, become memory B cells, or migrate to the dark zone, where they proliferate and mutate their antibody genes. The latter cells circulate back to the light zone and become subjected to the same selection process. Antibodies secreted by plasma cells can feedback into the GC and affect the selection process. (B) Schematic of the simulations. (Top) Timeline showing dose administration and corresponding antigen levels. (Bottom) GCs are formed following the prime and gradually shrink with time due to decreasing antigen levels. The prime could be low dose (LD) or standard dose (SD). The boost could restore existing GCs (mechanism I) or lead to new GCs seeded by memory B cells formed during the prime (mechanism II). The boost is typically SD.

Following dosing, antigen is trafficked to the lymph nodes, where its levels rise rapidly and then decline exponentially (34, 46). Accordingly, we let η rise immediately upon dosing to a pre-determined amount dependent on the vaccine dosage and then decrease with each generation based on the half-life of the administered antigen (Fig. 1B). With the boost, we considered two scenarios (34, 47, 48): the first where the boost enhanced antigen levels in pre-existing GCs, and the second where it initiated new GCs using memory B cells formed by the prime. We also examined the baseline, control scenario where the boost initiated GCs *de novo*, independently of the prime. We considered vaccination protocols with low and standard dose prime and a range of prime-boost dosing intervals. We performed multiple stochastic realizations of the simulations for each vaccination protocol and predicted the expected antibody response as an indicator of vaccine efficacy.

### Antigen availability and its effect on selection stringency

To elucidate affinity maturation in the GC reaction, we first performed simulations with a constant η, set here to 7. (We considered other values of η later.) The GC initially had B cells with low affinity for the target antigen. As the GC reaction proceeded, B cells with increasing affinity were selected in our simulations, marking affinity maturation (Fig. 2A). Eventually, a stationary distribution of B cells of different affinities was achieved, dominated by B cells with the highest affinities, as observed in previous studies (39) and akin to the mutation-selection balance observed in other evolutionary simulations (49, 50). We focussed on the corresponding evolution of the average affinity of the B cells. As the GC reaction progressed, the average affinity of the B cells increased and reached a plateau (Fig. 2B). Thus, when η=7, the average affinity of the B cells, determined by the average match-length between the antigen and BCR sequences, plateaued at ∼6.7 (Fig. 2B inset). Note that *L*=8 in these simulations.

**Figure 2.**
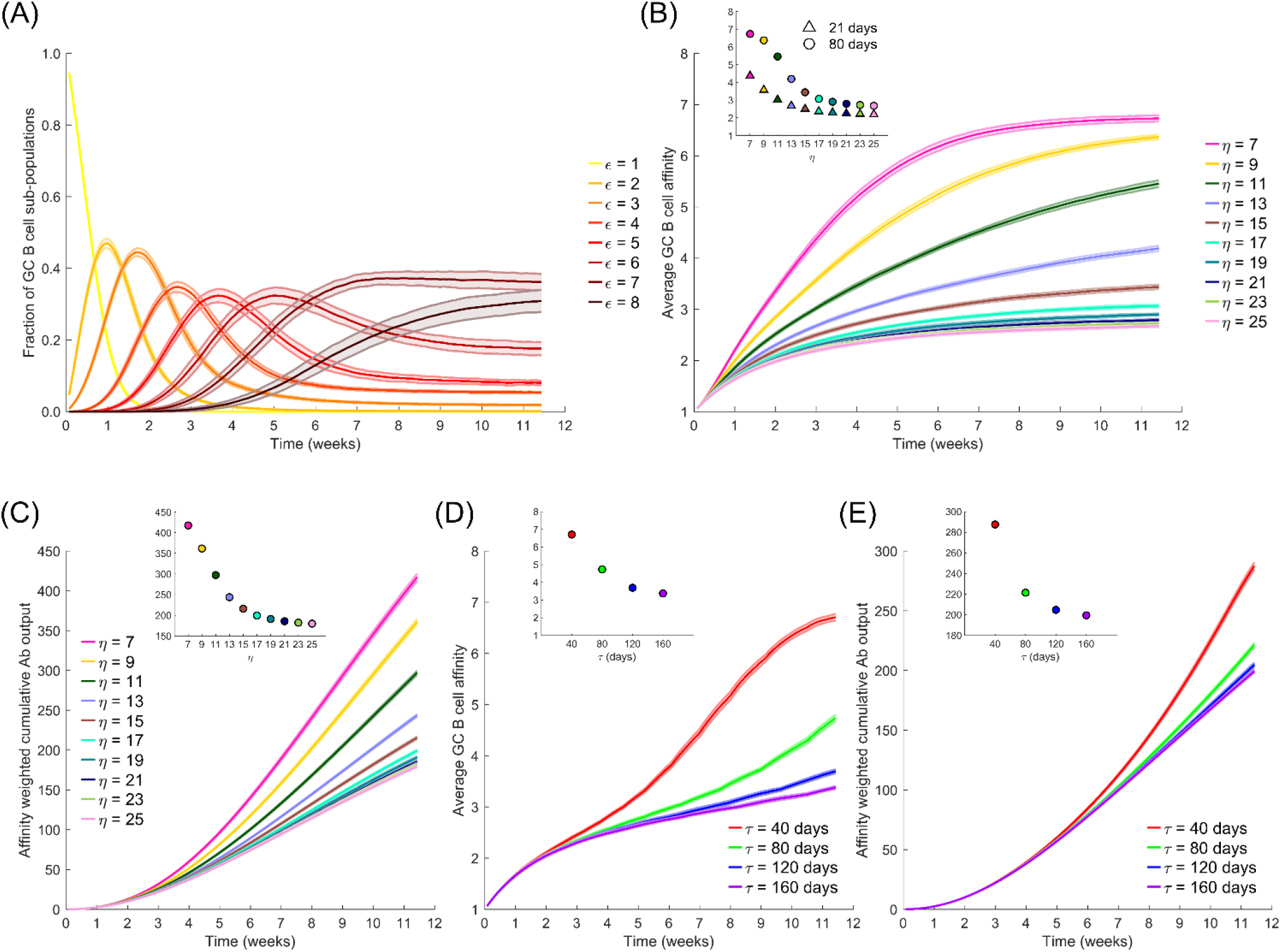
The effect of antigen availability and half-life on the GC reaction. (A) Time-evolution of populations of GC B cells of different affinity, ε, for the antigen; η=7. (B) Time-evolution of the average affinity of GC B cells for different η. *Inset*: The values at day 21 and day 80, the latter the plateau values, of the average affinity versus η. (C) Time-evolution of the affinity-weighted cumulative antibody output for different η. (D) Time-evolution of the average affinity for different antigen half-lives, τ, and the initial η, η_0_=20. (E) Corresponding cumulative antibody output. *Insets in* (*C-E*): Corresponding values at day 80.

To examine the effect of antigen availability, we next performed simulations at different values of η. Increasing η resulted in a lower value of the plateau of the average affinity (Fig. 2B), indicative of weaker selection. Increasing η would correspond to higher vaccine dosages. B cells with lower affinities were selected with higher η because more opportunities were available for antigen acquisition. Thus, the average affinity plateaued at ∼3.4 when η=15 and decreased further with larger η (Fig. 2B inset). This is consistent with the classic observations of poorer affinity maturation with increasing antigen levels (26, 39). In terms of the absolute antibody titres, our simulations predicted that unless the selection stringency was so large that the GC B cell population began to decline causing GC collapse (Fig. S1), the GC B cell population was maintained, leading to a steady output of Abs from the GC (Fig. S2). The lower affinity with increasing η thus resulted in a corresponding decrease in the affinity-weighted cumulative antibody output in our simulations (Fig. 2C). The latter output was ∼417 when η=7 and ∼216 when η=15 at 80 d following dosing (Fig. 2C inset). This affinity-weighted antibody output would serve as a measure of the humoral response elicited by vaccination; it accounts for the effects of both the quality and the quantity of the response. At very high values of η, beyond ∼20 in our simulations, the effect of varying η was minimal (Fig. 2B and C), indicating that at sufficiently high dosages, the effect of varying dosage on the GC reaction may not be significant. At lower η, between 7 and 15 in our simulations, lowering dosage resulted in a substantial gain in the GC response. When η was too low, however, in our simulations, GCs collapsed, as not enough antigen was available for sustaining the B cell population (Fig. S1).

Following vaccination, antigen levels are expected to decline exponentially with time. We therefore next performed simulations with η decreasing with a half-life τ; i.e., η=η_0_exp(-t×ln2/τ), where η_0_ is the peak antigen level achieved soon after dosing. How antigen levels quantitatively decay on follicular dendritic cells within GCs relative to that in plasma is not well understood (34, 51, 52). We therefore examined a range of values of τ. We found in our simulations with η_0_=20, that the average affinity was higher when τ was lower (Fig. 2D). Specifically, the average affinity at day 80 from the start of the GC reaction was ∼6.7 for τ=40 d and ∼3.4 for τ=160 d (Fig. 2D inset). The faster decay of antigen thus increased the selection stringency within the GC and led to higher affinity B cells. The affinity-weighted cumulative antibody output, accordingly, increased with decreasing τ, consistent with an improved response due to increased selection stringency (Fig. 2E).

### Prime-boost vaccination: the effect of dosage

We now applied our simulations to mimic the prime-boost vaccination protocols employed in clinical trials (5). Specifically, we considered low dose (which we set using η_0_=10) and standard dose (η_0_=20) combinations, administered with a dosing interval Δ=28 d mimicking experimental protocols (5, 6, 21). (Our conclusions are not sensitive to these parameter settings; see Fig. S3) An important aspect of the humoral response associated with multiple antigen dosing that remains unknown is whether the subsequent doses modulate GCs formed following the first dose or seed new GCs. GCs have been observed to persist over extended durations following COVID-19 vaccination (24). (Such persistent GCs have been seen following natural infection with other viruses too (25).) If the interval Δ is relatively small, one may expect the boost to modulate ongoing GC reactions, as has been suggested previously (34, 39). However, if Δ is large, then the GCs formed by the prime may collapse due to antigen decay before the boost, so that the seeding of new GCs by the boost is more likely. In the latter scenario, the effect of the prime must come from the preferential seeding by memory B cells formed following the prime (47, 48, 53). Recruitment of memory B cells into GCs has been suggested, especially those B cells that displayed cross reactivity to other circulating human betacoronaviruses (24). We therefore simulated two limiting scenarios (Fig. 1B): First, we assumed that the boost modulated existing GCs and seeded no new GCs. Second, we let the boost seed GCs using the memory B cells formed from the prime and not modulate any existing GCs. We also simulated a control case where the boost established new GCs *de novo*, without using memory B cells from the prime, in which case no advantage from the prime is expected.

With the boost modulating existing GCs, our simulations predicted an advantage of the low dose prime over the standard dose prime (blue and red lines in Fig. 3A, B). The average affinity increased with time more steeply with the low dose until day 28, when the boost was administered (Fig. 3A). Just prior to boost administration, the average affinity was ∼4.9 for the low dose versus ∼2.8 for the standard dose prime. Correspondingly, the affinity-weighted cumulative antibody output was higher for the low dose than the standard dose (Fig. 3B). The administration of the boost caused an increase in antigen availability (Fig. 3A inset), relieving the selection stringency. The average affinity thus saw a temporary dip (Fig. 3A). However, as affinity maturation continued, the higher affinity B cells selected with the low dose prime expanded substantially, yielding a much higher affinity-weighted antibody output than with the standard dose prime (Fig. 3B). The average affinity and the affinity-weighted cumulative antibody output was higher with the low dose prime than the standard dose prime throughout our simulations.

**Figure 3.**
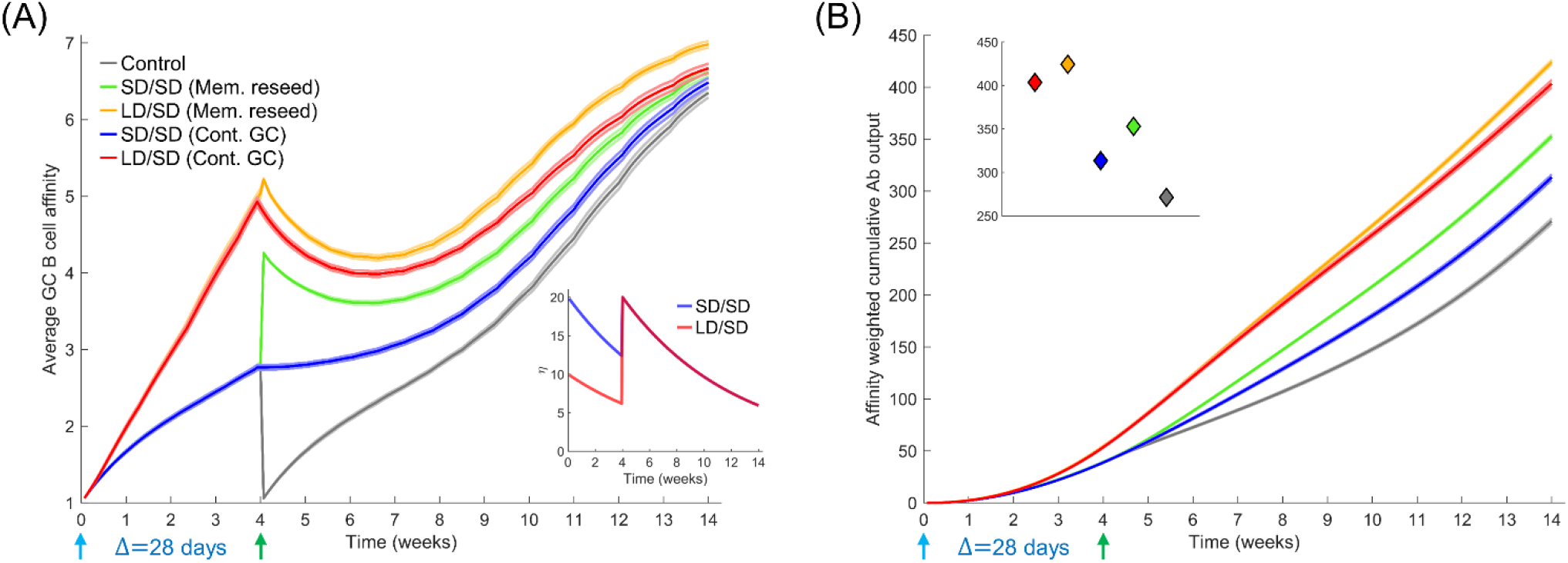
Influence of different prime-boost dosages. (A) Time-evolution of the average affinity of GC B cells for different dosing protocols indicated. *Inset*: The associated antigen levels. (B) Time-evolution of the affinity-weighted cumulative antibody output for the cases in (A). *Inset*: Corresponding values at the final simulation time point. Parameters used: Δ=28 d; τ=40 d; η_0_=10 for LD and η_0_=20 for SD.

When we let the boost seed GCs using memory B cells from the prime, the difference between low dose and standard dose prime was smaller in our simulations following the boost (green and orange curves in Fig. 3A, B). This is because we assumed that only B cells above a certain affinity for the antigen (here, match length ≥ 3; see Methods) could differentiate into memory B cells following stimulation. The advantage of the low dose prime in yielding high affinity B cells was thus reduced. The choice of memory B cells is in keeping with the expectation that low affinity naïve-like B cells may not receive strong enough signals to differentiate into switched memory B cells (54). Also, low affinity B cells are likely to exist regardless and thus seeding GCs with low affinity memory B cells may be no different from seeding GCs *de novo*. Yet, even within the memory pool, the low dose prime yielded higher affinity B cells than the standard dose prime, explaining the advantage of the low dose prime in our simulations (Fig. 3A). The differences in the corresponding affinity-weighted cumulative antibody output (Fig. 3B) were as expected but commensurately smaller than when the boost seeded existing GCs. Both scenarios yielded better responses than the control case where the boost seeded GCs *de novo* (grey lines in Fig. 3A, B).

### Prime-boost vaccination: the effect of dosing interval

To assess the influence of the dosing interval, we compared next the antibody responses elicited by two dosing intervals, Δ=28 d and Δ=56 d. We let τ=80 d here to avoid GC collapse following low dose prime with shorter antigen half-lives (Fig. S4). The average GC B cell affinity was significantly higher with Δ=56 d than Δ=28 d when the GCs were allowed to persist until the boost (Fig. 4A, B). For instance, the average affinity was ∼6.6 and ∼4.4, respectively, in the two cases, just before the administration of the boost following low dose prime, because affinity maturation continued longer with the longer dosing interval. Besides, the declining antigen levels further increased selection stringency in the latter case. This qualitative trend remained with the standard dose prime. The affinity-weighted cumulative antibody output was also significantly higher with the Δ=56 d than Δ=28 d (Fig. 4C, D). For instance, 28 d after the boost, the output was ∼380 and ∼174, respectively, in the two cases, when low dose prime was used and the boost modulated existing GCs. With standard dose prime too, the difference was nearly 2-fold. This effect remained whether the boost seeded new GCs or modulated surviving GCs (Fig. 4A-D), indicating a distinct advantage of the longer interval. The cases all yielded significantly better responses than the control case where the boost elicited GCs *de novo* (Fig. 4A-D).

**Figure 4.**
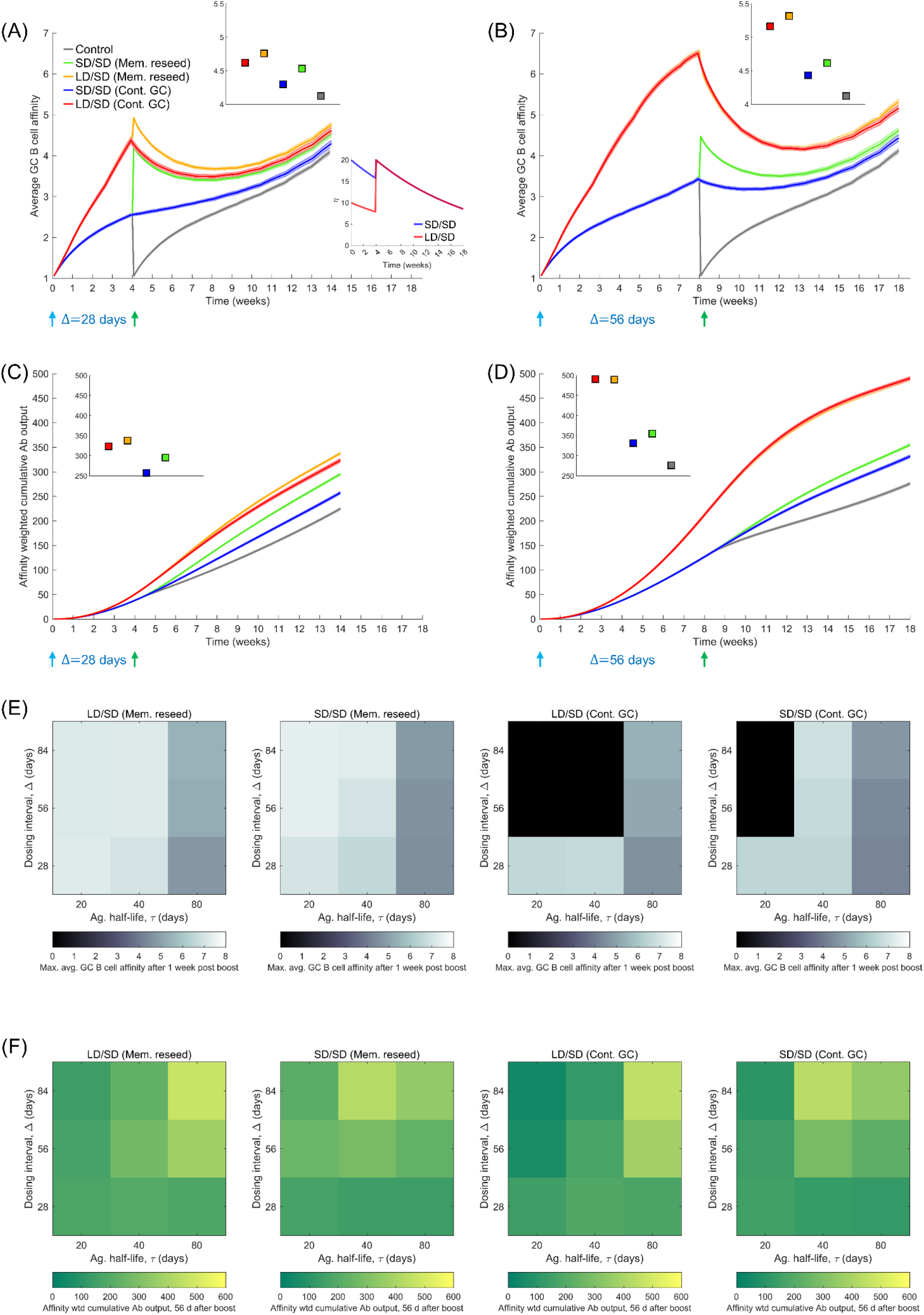
Influence of prime-boost dosing interval. (A, B) Average GC B cell affinities, and (C, D) affinity-weighted Ab outputs, with prime-boost intervals (Δ) of either 28 d (A, C) or 56 d (B, D), and with LD/SD or SD/SD dosing. Bottom inset of (A): LD and SD correspond to η_0_=10 and 20, respectively, with τ=80 d. Top insets in (A-D): values at the final time point. Heatmaps of (E) the maximum GC affinity recorded between 1 week post boost administration and the final time point, and (F) the affinity-weighted cumulative Ab output 28 d post the boost, as a function of τ (20, 40 and 80 d) and Δ (4, 8, and 12 weeks) for the two limiting scenarios (Mem. reseed and Cont. GC). Trajectories corresponding to the heatmaps are shown in Figure S4. Black regions in (E) correspond to collapsed GCs. A heatmap of the affinity-weighted cumulative Ab output 56 d post the boost is shown in Figure S4C.

Because the selection stringency depended on antigen half-life, τ, we assessed the effect of varying Δ for a range of values of τ. Following recent experiments (9, 12), we also considered much larger values of Δ; ranging from 28 d to 84 d (Figs. 4E, F and S4). To evaluate the effect on affinity maturation, we compared the maximum value of the average GC B cell affinity achieved at any time 1 week post the boost (to eliminate transients). We found that at any τ, increasing Δ increased the peak affinity, regardless of the use of low dose or standard dose prime or whether the boost seeded new GCs or affected existing GCs (Fig. 4E). Thus, a longer duration yielded a GC response of better quality. Further, the lower was τ, the higher was the peak affinity at any Δ, consistent with stronger selection stringency associated with lower antigen availability (Fig. 4E).

This latter effect influenced the overall response, combining quality with quantity, which we assessed using the affinity-weighted cumulative antibody output 28 d post the boost (Fig. 4F). While the overall trend of improved output with longer Δ remained, the trend was more nuanced. The nuances were due to the complex dynamics of the GC responses following multiple dosing. We examined first the effect of low dose prime. When τ was large, the GC reaction was sustained longer, allowing greater affinity maturation (Fig. S4). Thus, delayed dosing interval would lead to better responses. Indeed, with Δ=56 d and Δ=84 d, our simulations predicted that the cumulative output improved with τ (Fig. 4F). With Δ=28 d, the GCs may not have expanded sufficiently before the boost. With low τ, leading to high selection stringency, GCs tended to collapse after the boost (Fig. S4). With large τ, the selection stringency was weaker and it therefore took longer for affinities to rise. Consequently, intermediate τ yielded the best response (Fig. 4F).

With standard dose prime, too, the effects were similar. The GCs were sustained longer as τ increased, but weaker selection due to greater antigen availability led to poorer affinity maturation (Fig. S4). The trade-off tended to yield the best response at intermediate τ. In our simulations, when the boost contributed to existing GCs, it was not efficient in rescuing GCs that were beginning to collapse. Thus, with low and intermediate τ, GCs tended to collapse (Fig. S4). When the boost was assumed to seed new GCs using memory cells from the prime, because the latter had higher affinities for the antigen, the GCs not only survived, but also expanded. The benefit was amplified with delayed dosing as better memory cells became available for seeding the GCs. Thus, as long as τ was not too small, the cumulative output tended to improve with increasing Δ (see τ=40 d and 80 d in Fig. 4F). (With very small τ, the increased GC collapse compromised the response at high Δ; see τ=20 d in Fig. 4F). These trends were maintained when the output was considered 56 d post boost (Fig. S4). That GCs following COVID-19 vaccination can persist over extended durations (24) suggests that GC shrinkage may be slow *in vivo*. Large dosing intervals would then improve responses, as has been observed in clinical trials (9).

## DISCUSSION

Understanding the reasons behind the improved efficacy of COVID-19 vaccines upon delaying the boost dose or using a low dose prime would aid optimal deployment of vaccines, critical to settings with limited supplies. Here, using comprehensive stochastic simulations of the GC reaction post vaccination, we elucidated plausible mechanistic origins of the improved efficacy. To our knowledge, ours is the first study to employ such simulations to assess the influence of COVID-19 vaccination protocols. The GC reaction is constrained by a quality-quantity trade-off (26, 34, 35, 39): Lower antigen availability in the GC leads to more stringent B cell selection, resulting in the production of higher affinity antibodies but in smaller amounts. Increasing antigen availability reverses these effects. The different dosing protocols used–low versus standard dose prime and different dosing intervals–affect this trade-off. With low dose prime, antigen availability in the GCs is lowered, resulting in the selection of high affinity GC B cells. The boost relaxes the selection stringency and allows the expansion of the selected B cells. Delaying the boost delays the relaxation, resulting in even higher affinity B cells getting selected following the prime. Following the boost, these latter B cells would result in better overall GC responses, explaining the observed improvements in efficacy.

Experimental evidence supports the above reasons. Antibody titres targeting the SARS-CoV-2 spike were measured in individuals administered the boost 8-12 weeks, 15-25 weeks, and 44-45 weeks after the prime (9). The titres were consistently higher in the individuals with the longer dosing intervals. However, interestingly, the titres just before the boost, were lower in the individuals with the longer intervals. This was consistent with lower antibody output due to declining antigen availability with time in the GC and the associated GC shrinkage. Furthermore, the higher corresponding selection stringency may have resulted in the selection of GC B cells and memory B cells with higher affinity, which would be expected to rescue shrinking GCs or seed new GCs better, explaining the better responses eventually observed. Improved antibody responses following delayed boost dosing has now been observed with multiple vaccines (9-12).

With dosing intervals smaller than 8-12 weeks or with the low dose prime, the differences in antibody titres have been less apparent (5, 8, 21). Yet, the improvement in vaccine efficacy is substantial (5). While we have argued that this improvement may be due to the improved affinity of the antibodies, direct measurements of affinity are lacking. *In vitro* pseudo-typed virus neutralization efficiency of antibodies isolated 28 d after the boost were not significantly different between individuals administered the low dose prime or the standard dose prime or when both standard doses were administered with a 28 d or 56 d interval (5, 8, 21). It is possible that the improvements in affinity may not be adequate to be manifested as improved *in vitro* neutralization efficiencies, possibly because the stoichiometry of antibody binding to the viral spike proteins that ensures virus neutralization (55-57), which is yet to be estimated for SARS-CoV-2, may be realized in both scenarios. *In vitro* neutralization efficiencies tend to be much higher than corresponding *in vivo* efficiencies (58). Nonetheless, greater affinity maturation with lower antigen availability has been long recognized as a hallmark of the GC reaction (26, 34, 35, 39). In independent studies on HIV vaccination, for instance, protocols that allowed antigen levels to rise with time, akin to low dose prime followed by standard dose boost examined here, elicited better antibody responses than protocols that held the antigen levels constant or allowed them to decline with time (34), an effect consistent with the dosing protocols modulating antigen availability and the associated quality-quantity trade-off in the GCs (39).

Our simulations predicted a role for antigen half-life in the response to vaccination. With longer half-lives, the response improved upon increasing the dosing interval. With shorter half-lives, if associated GC shrinkage was too drastic before the administration of the boost, the response following the boost was compromised. Shorter dosing intervals then elicited the best response. We note here that the antigen half-life in the GC may be difficult to estimate (34, 51, 52). That GC B cells and plasmablasts were detectable in high frequencies even 12 weeks after the boost suggests that antigen presented by COVID-19 vaccines may be much longer lasting in the GCs than expected from their half-life in circulation (24). Such prolonged GC responses have been observed in other settings (25). Future studies may yield accurate estimates of the antigen half-life in GCs, which would help identify optimal dosing intervals for the different COVID-19 vaccines available.

Quantitative comparison of our predictions with experimental observations is difficult, as has been the case with other modeling studies of the GC reaction (34-36, 39, 40, 42, 45). This is because a number of key biological processes associated with the GC reaction remain to be elucidated, including the link between dosage and the number of GCs seeded, and between measurable antigen levels in circulation and those within individual GCs (22, 23, 34, 35, 39). Only recently have these links begun to be evaluated (37). As a simplification, our simulations have assumed that increased dosage leads to increased antigen availability within GCs while keeping the number of GCs seeded fixed. It is possible that the number of GCs seeded may also increase with dosage but with a commensurately smaller rise in the antigen levels per GC. Future studies that elucidate the links above may help define these quantities better. Nonetheless, the poorer quality of the antibody response with increasing dosage is a widely observed and accepted phenomenon (26, 34, 39), giving us confidence in our findings.

We recognize that other arms of the immune system that could be triggered by the vaccines, particularly T cells, may affect the vaccine efficacies realized (5-9, 13, 14). The strength and timing of the T cell response has been argued to be important in determining the severity of the infection (59), which in turn may affect the estimated vaccine efficacy (60). We have focused here on the antibody response, to which the efficacies have been found to be strongly correlated (18, 19, 60), and which in our simulations qualitatively explained the effects of the different dosing protocols on vaccine efficacies.

Other hypotheses have been proposed to explain the effects of low dose prime and delayed dosing intervals, the predominant of which has been the undesirable response to the adenoviral vector in the case of the Oxford/AstraZeneca vaccine that could blunt the response to the boost (61). While these hypotheses remain to be tested, that the effects are now evident with more than one vaccine, including lipid nanoparticle mRNA vaccines that do not use the adenoviral vectors (10-13), suggests that the effects are intrinsic to the responses elicited by the SARS-CoV-2 antigens in the vaccines, supporting our hypothesis.

In summary, our study offers an explanation of the confounding effects of different dosages and dosing protocols on COVID-19 vaccine efficacies. The resulting insights would inform studies aimed at designing optimal vaccine deployment strategies.

## METHODS

### Stochastic simulations of the GC reaction

We developed the following *in silico* stochastic simulation model of the GC reaction (Fig. 1A). The model builds on a previous study which examined the role of passive immunization on the GC reaction (39). Here, we adapted it describe the effect of COVID-19 vaccination.

### Initialization

We initiated the GC reaction with *N*=1000 GC B cells of low affinity for the target antigen in the light zone of the GC. This follows observations where low affinity seeder B cells initiate the GC reaction by proliferating rapidly to a steady state size of 1000 cells, following which somatic hypermutation and affinity maturation commence (36, 39). We considered a non-mutating antigen, determined by a randomly chosen string of length *L* and alphabet of size *к*=4. The alphabet size represents the broad classes of amino acids, namely, positively charged, negatively charged, polar, and hydrophobic (42). The B cell receptor (BCR) paratope for each cell is then set by randomly mutating the antigen sequence at *L*-1 randomly chosen positions. This ensured that the cells in the initial pool all had low affinities for the antigen. The B cells were then allowed to acquire antigen.

### Antigen acquisition

Antigen is presented to B cells as antibody-bound immune complexes on follicular dendritic cell surfaces. The probability with which a B cell successfully acquired the antigen was *f*_*Ag*_ = (*ε* − *ω* + *L*) / 2*L*, where ε and ω are the lengths of the longest common substrings of the antigen sequence and those of the associated B cell receptor (BCR) and the presenting antibody, respectively. The latter expression followed from a mechanistic consideration of bond dissociation triggered by the competition between the BCR and the antibody for the antigen (39). Note that antibodies are secreted versions of the BCRs and hence were similarly represented as bit-strings of length *L* too. The presenting antibodies were produced by plasma cells and re-entered the GC via antibody feedback, described below. B cells were selected at random for antigen acquisition, with each B cell selected η times on average. The amount of antigen acquired by a B cell was set equal to the number of successful acquisition attempts, denoted as θ. B cells had to acquire a minimum amount of antigen, denoted θ_c_, for them to survive. Surviving cells were eligible to receive help from T follicular helper (T_fh_) cells. We capped the level of antigen acquired at θ_∞_, at which point the B cell may have received saturating levels of stimulatory signals necessary for T_fh_ cell help.

### T_fh_ cell help

We chose surviving B cells randomly and let each cell receive T_fh_ cell help with the probability *f*_*T*_ = (*θ* −*θ*_min_) / (*θ*_max_ −*θ*_min_), where θ_min_ is the minimum antigen acquired by the surviving B cells and θ_max_ (= min (*η,θ*_∞_)) is the maximum antigen acquired. The probability follows from the recognition that T_fh_ cell help depends on the relative and not absolute amount of antigen acquired (23, 39). Cells that do not receive help died. We continued this with every surviving cell and stopped if 250 cells successfully received T_fh_ help.

### Cell fate decision

Of the cells selected above, we chose 5% randomly to become memory B cells; 5% to become plasma cells; and the rest to migrate to the dark zone of the GC. The memory B cells were constrained to have a minimum affinity for the antigen (54) (here, match length 3) and were allowed to survive long-term. The plasma cells exited the GC, commenced producing antibodies, and died at the rate of 0.015 per generation (Table S1).

### Proliferation and mutation

The cells in the dark zone were allowed to multiply, with each cell dividing twice. Of the resulting cells, we chose 10% and introduced single random point mutations in their BCR sequences. The latter frequency was chosen following estimates based on the somatic hypermutation frequency suggesting that 1 in 10 GC B cells would be mutated per generation in their antibody variable region genes (23, 36, 39, 42). The two divisions per cell would bring the cell population back to the N∼1000 cells. This completed one generation of B cell evolution in the GC.

### Recycling

The resulting cells in the dark zone were all allowed to migrate to the light zone, offering the next generation of cells on which the above process would repeat.

### Antibody feedback

Antibodies produced by plasma cells could traffic back to the GC and influence antigen presentation (35). Accordingly, following estimated trafficking timescales, we let antibodies produced by plasma cells in any generation become the antibodies presenting antigen to B cells two generations later (35, 39). Antibodies were also systemically cleared at the rate of 0.01165 per generation (Table S1).

### Termination

We repeated the above process for up to 250 generations (∼18 weeks) or if the cell population declined, leading to GC collapse.

### Dosing protocol

We implemented the prime-boost dosing protocol by letting η vary with time as η=η_0_exp(-t×ln2/τ), mimicking antigen rise immediately upon dosing (to η_0_) and an exponential decline subsequently with half-life τ (34, 39, 46). The decline is assumed to subsume any loss of antigen due to acquisition by B cells. We set η_0_ based on whether a low or standard dose was employed. The prime and boost were separated by the duration Δ. Our interest is in large values of Δ and low first dosages, so that at the time of boost administration, the residual antigen is small. Whether memory B cells seed GCs post boost is a topic of active current research (47, 48, 53, 54, 62, 63). We therefore considered all potential scenarios, with the boost 1) feeding into existing GCs; 2) seeding new GCs using memory B cells; 3) seeding new GCs using naïve B cells. In scenario 2, we let the memory B cells for seeding the GCs be chosen with a probability proportional to their affinity for the antigen. In other words, the distribution of B cells of different affinities in the seeder pool mimics the distribution of affinity-weighted fractions of memory B cells formed following the prime.

### Parameter values

The parameter values employed and their sources are listed in Table S1.

### Quantification of the GC response

With each parameter setting, we performed 2500 realizations, which we divided into 25 ensembles of 100 GC realizations each (39). The average GC B cell affinity in the *g*^th^ generation was calculated using,

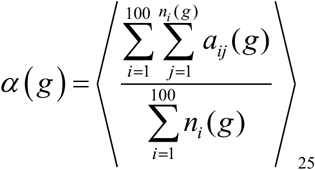

where *a*_*ij*_ was the affinity of the *j*^th^ B cell among the *n*_*i*_(*g*) B cells in the *g*^th^ generation of the *i*^th^ realization of an ensemble. The angular brackets represent averaging across the ensembles. The affinity-weighted plasma cell output in the *g*^th^ generation was

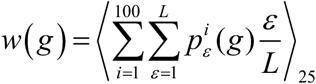

where 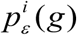 was the number of plasma cells with affinity ε in the *g*^th^ generation. If plasma cells died at the per capita rate δ_p_, then the affinity-weighted cumulative plasma cell output would be

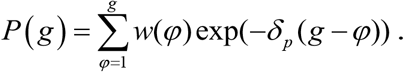

If the antibody production rate of plasma cells was β per generation (64), the instantaneous affinity-weighted antibody output would be βP(g), which given the clearance rate, δ_A_, of circulating antibodies yielded the affinity-weighted cumulative antibody output as

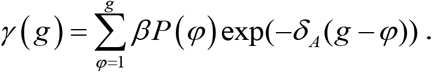

We performed the simulations and analysed the results using programs written in MATLAB.

## Data Availability

All data is available within the manuscript and supplementary information.

## Supplementary Materials for

**Figure S1.**
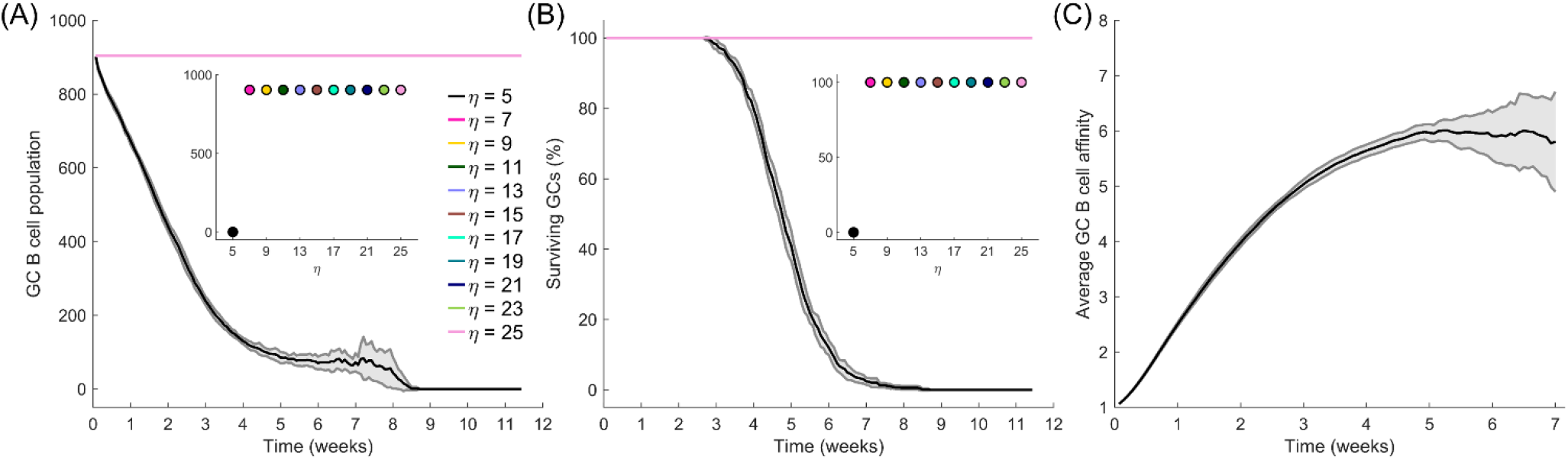
Influence of η. Time series of (A) GC B cell population and (B) percent surviving GCs for simulations with various η. GCs with low η (=5 here) have high rates of apoptosis which result in GCs gradually being extinguished with time. However, these GCs also exhibit accelerated affinity maturation (C) due to the higher selection stringencies. Insets: values at the final time point.

**Figure S2.**
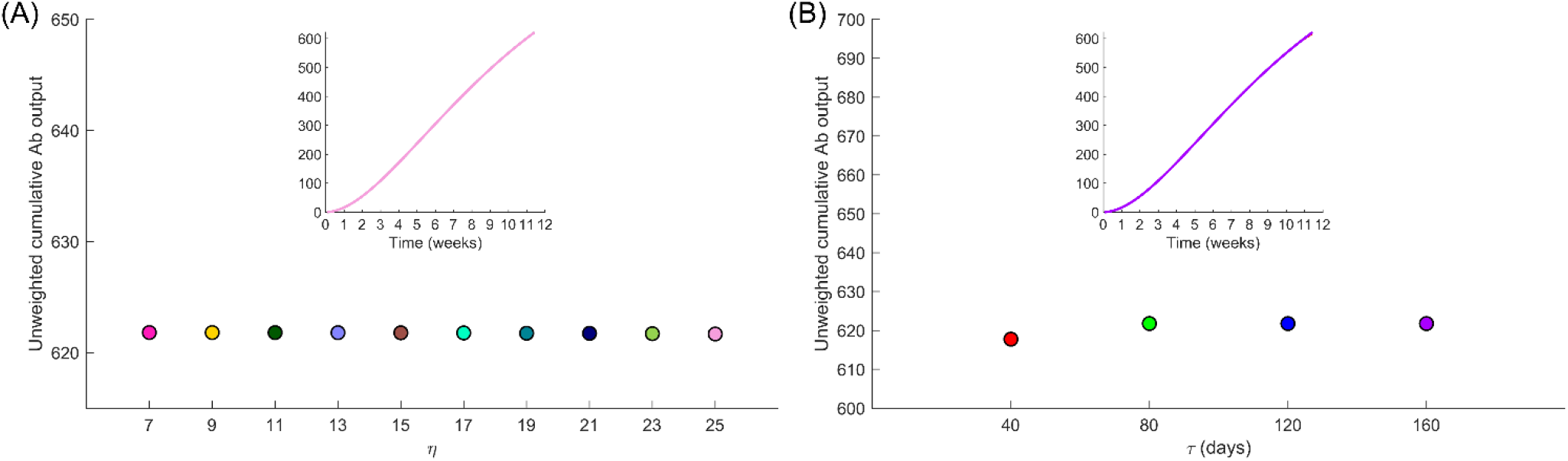
Absolute Ab output. Unweighted cumulative antibody output at the final time point for various (A) η, and (B) τ. Insets: time series. The curves for the different cases overlap.

**Figure S3.**
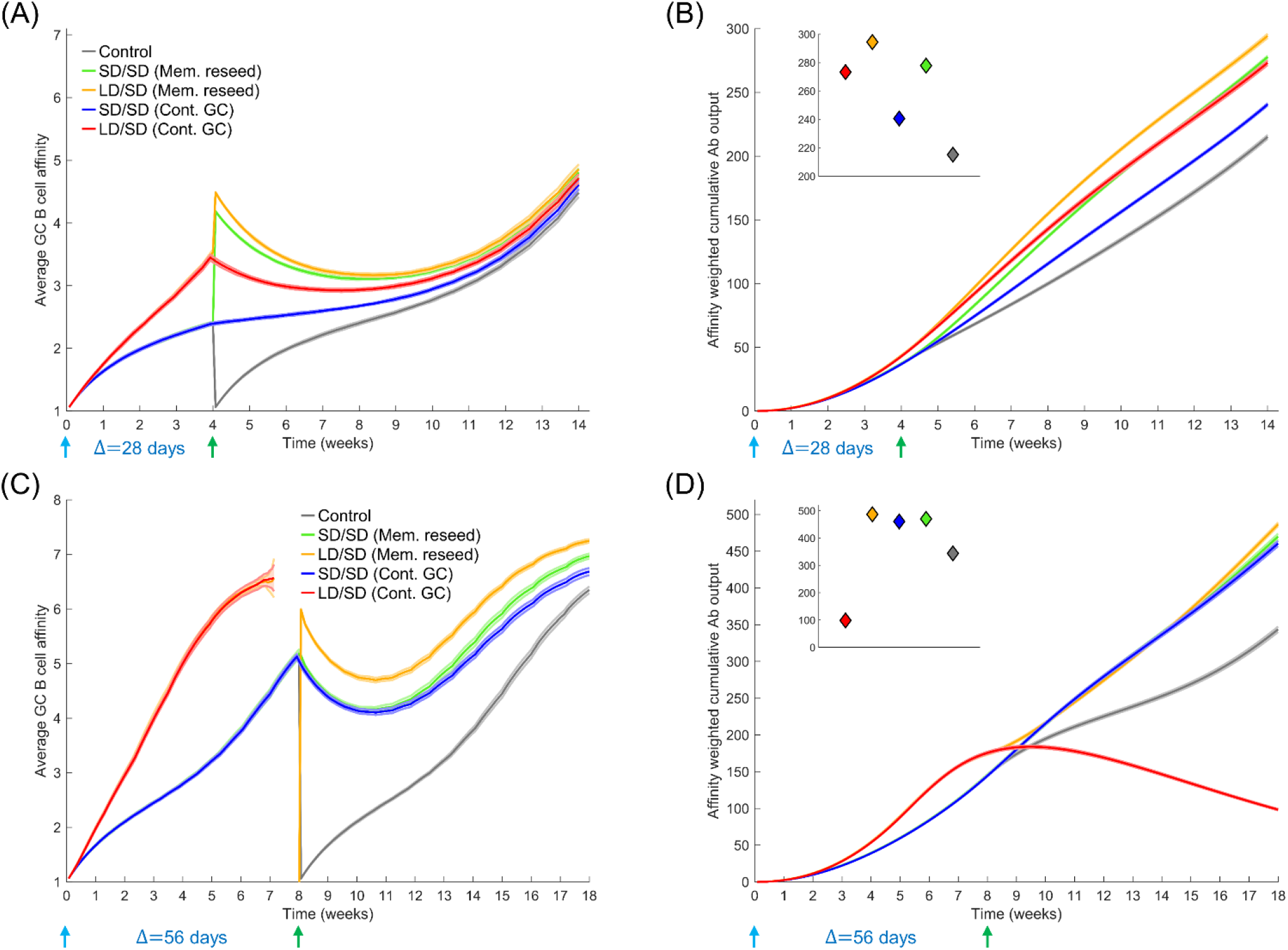
Influence of initial Ag levels (dose) and the prime-boost dosing interval (Δ). (A) Average GC B cell affinity, and (B) affinity-weighted Ab output with a prime-boost interval Δ=28 d and antigen half-life τ=40 d, either with LD/SD or SD/SD dosing (LD and SD correspond to η_0_=15 and 30, respectively). These trends are qualitatively similar to those in Figure 3. (C) Average GC B cell affinity, and (D) affinity-weighted Ab output with Δ=56 d and τ=40 d, either with LD/SD or SD/SD dosing (LD and SD correspond to the default η_0_=10 and 20, respectively). Insets: values at the final time point.

**Figure S4.**
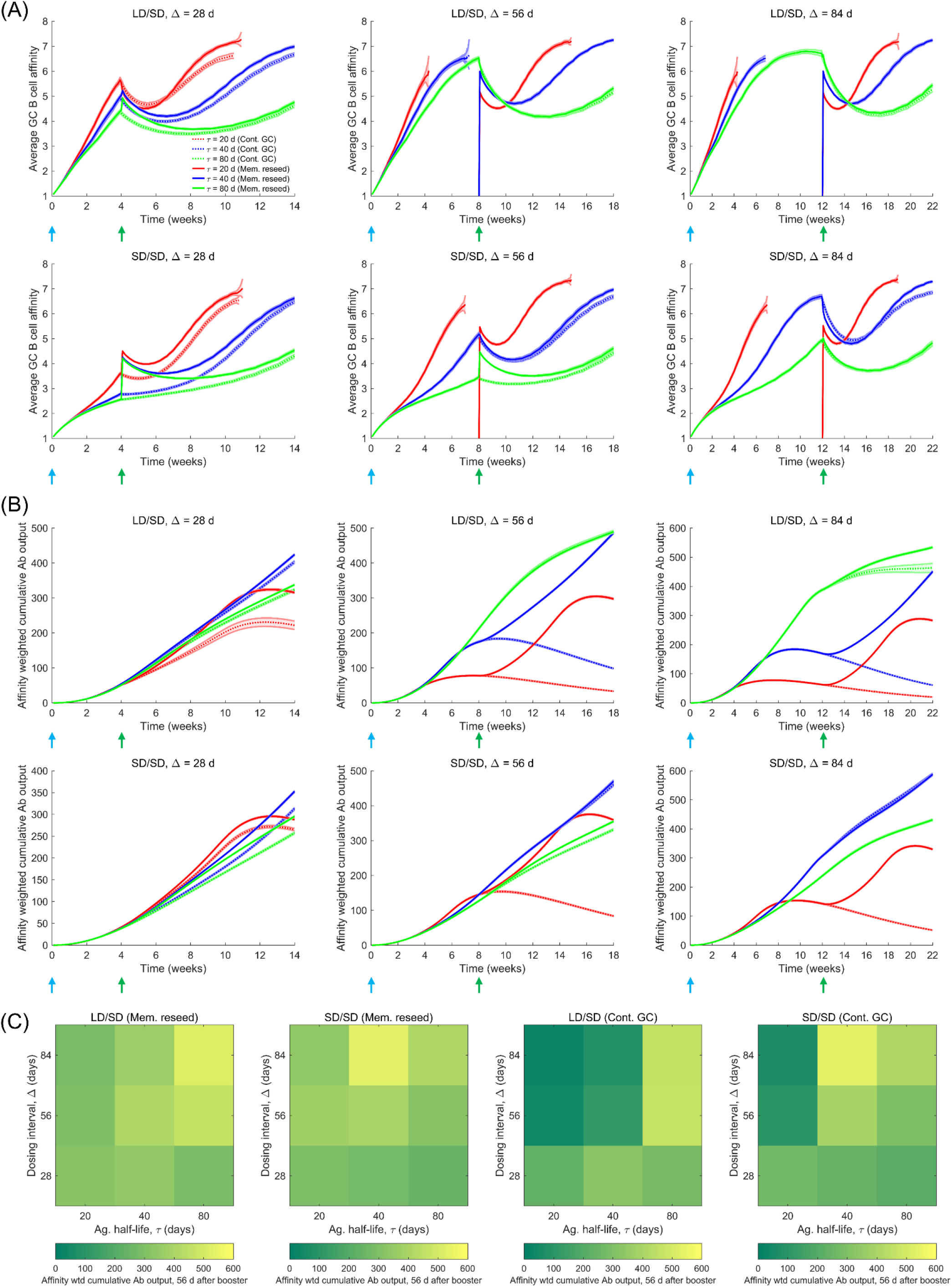
Influence of the prime-boost dosing interval (Δ). Time series of (A) average GC B cell affinities, and (B) affinity-weighted Ab outputs corresponding to the heatmaps in Figure 4E and 4F, respectively. (C) Heatmaps of the affinity-weighted cumulative Ab output 56 d post the boost, as a function of τ (20, 40 and 80 d) and Δ (4, 8, and 12 weeks) for the two limiting scenarios (Mem. reseed and Cont. GC).

**Table S1.** Model parameters and their values.

| *Symbol* | *Description* | *Value (units)* | *Refs.* |
| --- | --- | --- | --- |
| N | Number of B cells initiating the GC reaction | 1000 (cells) | (36, 39) |
| L | String length of antigen, B cell receptor (BCR), and antibody | 8 (dimensionless) | (39, 42) |
| $\kappa$ | Alphabet size for strings | 4 (dimensionless) | (39, 42) |
| $\theta_c$ | Minimum amount of acquired antigen for B cell survival | 3 (dimensionless) | (39) |
| $\theta_\infty$ | Maximum amount of antigen that can be acquired by a B cell | 5 (dimensionless) | - |
| $\delta_p$ | Clearance rate of plasma cells | 0.015 (generation<sup>-1</sup>) | (34, 44) |
| $\beta$ | Antibody production rate of plasma cells per generation | 2000 (molecules cell<sup>-1</sup> s<sup>-1</sup>) | (64) |
| $\delta_a$ | Clearance rate of antibodies | 0.01165 (generation<sup>-1</sup>) | (34, 44) |

